# Longitudinal Modeling of Intraocular Pressure and Its Associated Factors Among Glaucoma Patients: Evidence from Jimma University Medical Center

**DOI:** 10.64898/2026.10.20.26366063

**Authors:** Bikila Tegegn, Najmeh Nakhaeirad, Jaleta Abdisa, Meskerem Getachew, Tolesa Futesa Begna, Denekew Bitew Belay, Shiferaw Befkadu Bizuayehu

**Affiliations:** Department of Statistics, College of Natural Science, Jimma University, Jimma, Jimma, 378, Oromia, Ethiopia; Department of Statistics, University of Pretoria, Hatfield, Pretoria, 0028, Gauteng, South Africa; School of Health Systems and Public Health, Faculty of Health Sciences, University of Pretoria, Prinshof, Pretoria, 0084, Gauteng, South Africa; Department of Statistics, College of Science, Bahir Dar University, Bahir Dar, Bahir Dar, 79, Amhara, Ethiopia

**Keywords:** Glaucoma, Intraocular pressure, Longitudinal analysis, linear mixed effect model, factors, Bivariate analysis, Ethiopia

## Abstract

**Background:** Glaucoma remains one of the leading causes of irreversible blindness worldwide, and elevated intraocular pressure (IOP) is an important modifiable risk factor for disease progression. Monitoring changes in IOP over time is therefore essential for glaucoma management. However, evidence on the longitudinal trajectory of IOP and its associated factors remains limited in resource-limited settings. This study investigated the longitudinal trajectory of IOP and its associated demographic, socioeconomic, and clinical determinants among glaucoma patients at Jimma University Medical Center.

**Objective:** To assess the longitudinal trajectory of IOP and identify demographic, socioeconomic, and clinical factors associated with changes in IOP among glaucoma patients at Jimma University Medical Center.

**Method:** A retrospective longitudinal study was conducted among glaucoma patients who had at least three follow-up visits at Jimma University Medical Center. Repeated IOP measurements were obtained over six follow-up waves from January 2022 to January 2024. Profile plots were used to describe the longitudinal pattern of IOP, and linear mixed-effects models were fitted to assess factors associated with baseline IOP and changes in IOP over time. Separate models were fitted for the left and right eyes, in addition to a bivariate model that jointly accounted for both eyes.

**Results:** IOP generally declined over follow-up period, although the magnitude and rate of decline varied across patient characteristics and treatment groups. In the right Eye, rural residence was associated with lower baseline IOP, while occupation and treatment regimen were also significantly associated with baseline IOP. Significant time-by-occupation and time-by-treatment interactions were observed. In particular, right-eye IOP declined more rapidly among patients receiving Timolol with Diamox and Timolol with Xolamol. In the left eye, sex, occupation, treatment regimen, and treatment duration were associated with baseline IOP, while significant interactions between time and Timolol with Latanoprost and short treatment duration were observed. A bivariate model similarly showed significant effects of residence, occupation, and treatment regimen, as well as significant time-by-treatment interactions, indicating different longitudinal IOP trajectories across treatment groups.

**Conclusion:** IOP trajectories differed between the two eyes, with a significant decline in left-eye IOP but no significant overall temporal change in right-eye IOP. Treatment regimen was the most consistent factor associated with differences in the longitudinal trajectory of IOP, while occupation, residence, and treatment duration also showed significant associations in selected analyses. These findings highlight the importance of longitudinal monitoring and consideration of patient and treatment characteristics when assessing IOP among glaucoma patients. Further longitudinal studies with larger and more diverse populations are warranted to investigate additional behavioral, environmental, and clinical factors influencing IOP trajectories.

## 1 Introduction

Glaucoma is a chronic, progressive optic neuropathy and remains one of the leading causes of irreversible blindness worldwide. It is characterized by the gradual loss of retinal ganglion cells and optic nerve damage leading to irreversible visual field loss if it left untreated [1–3]. Although glaucoma may occur in individuals with normal intraocular pressure (IOP), elevated IOP remains the most important modifiable risk factor for disease onset and progression. Lowering IOP is therefore the primary treatment strategy for slowing disease progression and preserving vision [4–6].

The epidemiology of glaucoma continues to increase with population aging and longer life expectancy. Because the disease often develops without noticeable symptoms, many patients are diagnosed only after substantial and irreversible vision loss has occurred [2, 7]. Recent evidence indicates that glaucoma remains a major public health concern in Africa which has one of the highest regional burdens worldwide [8]. Vision is typically lost first in the periphery, producing visual field defects that respect the horizontal midline. As a result, glaucoma places a considerable burden on patients, families, and healthcare systems through lifelong treatment, regular follow-up, reduced quality of life, and loss of productivity, particularly in low- and middle-income countries like Ethiopia where access to specialized eye care is limited [2, 5]. The condition significantly impairs quality of life, with affected individuals especially those in developing countries, experiencing reduced functional status compared to non-glaucomatous peers. The constant fear of blindness and the demands of lifelong therapy can cause substantial physical and emotional burden [9].

Contemporary studies have shown that IOP is not a static clinical measure but changes over time. Longitudinal changes and fluctuations in IOP provide more useful information for assessing the burden of disease progression and treatment response than a single clinic measurement [7, 10, 11]. Moreover, recent research continues to support individualized IOP reduction as one of the most effective strategies for delaying glaucoma progression [12]. These findings highlight the importance of identifying factors that influence IOP during long-term follow-up.

Glaucoma progression and management is influenced by a combination of demographic, socioeconomic, and clinical factors. Age, place of residence, occupation, treatment duration, and treatment regimen may all affect long-term IOP control and disease progression [1, 4]. Patients living in rural areas are often experience delayed diagnosis and limited access to specialized eye care, while socioeconomic conditions and occupational demands may influence treatment adherence and continuity of care [1, 2]. Longer treatment duration, particularly with IOP-lowering medications, supports effective disease management and helps preserve optic nerve function, whereas inadequate or interrupted treatment contributes to further damage [6, 13]. These challenges are particularly important in Ethiopia, where glaucoma is a major cause of irreversible blindness and many patients present with advanced disease because of delayed diagnosis, limited awareness, and restricted access to specialized ophthalmic services, particularly outside major referral hospitals [14–16]. Treatment adherence and continuity of follow-up remain significant challenges in glaucoma care and may contribute to inadequate long-term disease control. Previous studies in Ethiopia have reported substantial difficulties with adherence to glaucoma treatment, with factors such as financial constraints, difficulties attending follow-up appointments, and longer intervals between visits contributing to poor adherence [16, 17]. Jimma University Medical Center is one of the largest teaching and referral hospitals in southwestern Ethiopia and provides comprehensive glaucoma care to a large catchment population from both urban and rural communities through specialized outpatient, diagnostic, laser, and follow-up services. Given the large and diverse population receiving care at Jimma University Medical Center, examining longitudinal changes in intraocular pressure (IOP) is important for understanding treatment response and disease control over time. Such information can support better patient management, guide treatment decisions, and contribute to evidence-based glaucoma care in Ethiopia.

Repeated IOP measurements collected from the same patient over multiple follow-up visits are naturally correlated and cannot be adequately analyzed using conventional regression methods that assume independent observations. Linear mixed-effects models provide an appropriate framework for analyzing longitudinal data by accounting for both the correlation among repeated measurements within patients and the variability between patients [18]. Although this approach has become increasingly common in longitudinal medical research, its application to glaucoma follow-up data in Ethiopia remains limited [7]. Addressing this methodological gap may provide more reliable evidence on longitudinal changes in IOP and its associated factors, ultimately supporting better clinical decision-making and glaucoma management.

Therefore, this study used a linear mixed-effect model to examine the longitudinal pattern of intraocular pressure among glaucoma patients attending Jimma University Medical Center. Specifically, the study assessed how demographic, socioeconomic, and clinical factors influence changes on IOP trajectory over time while accounting for the correlation among repeated measurements within patients. The findings are expected to improve understanding of IOP trajectory, support more individualized glaucoma care, strengthens patient follow-up, and contribute to the improvement of eye health services in Ethiopia and other resource-limited settings.

## 2 Data and Methods

This study was conducted at the Eye Clinic of Jimma University Medical Center (JUMC), located in Jimma, the capital of Jimma Zone in the Oromia Region of Ethiopia. Jimma is situated approximately 352km South-west of Addis Ababa. The study employed a retrospective longitudinal design using data from glaucoma patients who attended the Eye Clinic between January 2022 and January 2024. Patients who had at least three follow-up visits during the study period were included in the analysis. Repeated longitudinal measurements were obtained from patients’ medical charts across six follow-up waves occurring at approximately two-month intervals. Data were extracted from patient charts, which contained relevant epidemiological and clinical information. Socio-demographic informations were obtained directly from the patients during their follow-up visits. Patients’ medical charts were retrieved using registration card numbers obtained from the hospital’s electronic database system. Trained healthcare providers systematically extracted and documented the relevant socio-demographic and clinical information from the medical charts using the standard data extraction process.

### Source of Data and Quality Assurance

Patients were eligible in the study if they had attended the Eye Clinic for glaucoma management at least three times from January 2022 to January 2024. Patients with intermittent missing observations or loss to follow-up were retained in the study provided that they had at least three recorded visits. Missing data were handled using a multiple imputation procedure to obtain a complete longitudinal dataset for the analysis. Following imputation, the final dataset comprised 276 patients, each with six follow-up measurements, yielding a total of 1,656 patient-wave observations. Thus, the 1,656 observations represent the completed dataset after multiple imputation and do not imply that all patients had complete measurements at all six follow-up waves in the original medical records. Data quality was ensured through systematic data extraction and supervision by designated data controllers from the ophthalmic clinic.

### Inclusion and Exclusion Criteria

We included participants in the analysis if they attended at least three clinical visits, with data collected across six repeated waves. Participants could still be included even if they had intermittent missing data, dropped out, or were lost to follow-up during the study period. We excluded participants if they refused to participate, missed all previous measurement visits (i.e., were subject to attrition), or had fewer than three recorded visits during the study period. We also did not consider glaucoma patients who fell below the minimum data collection threshold in the final analysis.

### Variables of the Study

The response variable of this study was intraocular pressure (IOP) in millimetres of mercury (mmHg) for glaucoma patients. Changes in IOP over time were assessed and analyzed using a longitudinal approach.

The potential risk factors were categorized as

Socio-demographic factors: Age, gender, occupation, residence, alcohol, and smoking status;

Clinical factors: Surgical Interventions, types of treatment (e.g. specific medications or combinations), duration of treatment, blood pressure, and family history of glaucoma (genetic/hereditary predisposition).

### Statistical Analysis

The linear mixed model in longitudinal data analysis combine fixed effects to describe population-level associations with random effect to account for patient-specific differences and correlations among repeated measurements within patients. The general linear mixed effect model is defined as [19]

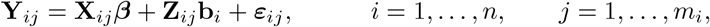

where **Y**_*ij*_ is the intraocular pressure (mmHg) for patient *i* at visit *j*, ***β*** represents fixed effects describing population-level associations, **b**_*i*_ represents patient-specific random effects, and ***ε***_*ij*_ accounts for within-patient variability. Model fitting involved estimating both fixed effects and variance components, including covariance parameters for random effects and residuals. Parameters in the linear mixed-effects models were estimated using Restricted Maximum Likelihood (REML), while Maximum Likelihood (ML) was used where appropriate for model comparison.

Potential explanatory variables were identified based on their clinical relevance and findings from previous studies. Before fitting the longitudinal model, the candidate variables were first screened using a p-value of 20% (*p <* 0.20). Variables that met this criterion, along with those considered clinically important, were included in the longitudinal linear mixed-effects model. A purposeful selection approach was then used to determine the variables retained in the final model, taking into account both their statistical contribution and clinical relevance.

Different linear mixed-effects models were compared using the Akaike Information Criterion (AIC), and Bayesian Information Criterion (BIC). The model with smaller AIC and BIC values was considered to have a better fit while maintaining model parsimony. The final model was chose based on these criteria, together with the clinical relevance and interpretability of the model.

The assumptions of the linear mixed-effects model were assessed using residual and Q-Q plots to evaluate homoscedasticity and normality, respectively. Potential outliers and influential observations were also examined using diagnostic plots, and multicollinearity among explanatory variables was assessed using the variance inflation factor (VIF). All data management, statistical analyses, model fitting, variable selection, and diagnostic checks were performed using R statistical software, version 4.5.3 (R Core Team, 2026, https://www.R-project.org)

### Ethical Considerations

Ethical approval for the study was obtained from the Research and Ethical Review Board (RERB/IRB) of Jimma University (Ref. No: RPG/184/2024). All patient information was treated as confidential and de-identified using unique codes. Physical records were securely stored in locked cabinets, while electronic data were kept on password-protected computers. Access to the data was limited to authorized members of the research team, and no personal identifiers were included in any report or publication.

## 3 Results

### Data Descriptions and Descriptive Statistics

Descriptive statistics for the socio-demographic, behavioral, clinical, and treatment-related characteristics are presented in Table 1. The study included 276 glaucoma patients who attended six follow-up visits at the Eye Clinic of Jimma University Medical Center between January 2022 and January 2024, contributing a total of 1,656 visits. Of the 276 patients, 151(54.7%) were male and 125(45.3%) were female, while 148(53.6%) resided in rural areas and 128(46.4%) in urban areas. Regarding occupation, 80 (29.0%) were civil servants, 58 (21.0%) were farmers, and 43 (15.6%) were daily worker.

**Table 1.** Descriptive Statistics of potential predictor variables for Categorical Variables.

| Variable | Category | Frequency | Percentage |
| --- | --- | --- | --- |
| Sex of patients | Female | 125 | 45.3 |
|  | Male | 151 | 54.7 |
| Residence | Urban | 128 | 46.4 |
|  | Rural | 148 | 53.6 |
| Occupation | Civil Servant | 80 | 29.0 |
|  | Merchant | 50 | 18.1 |
|  | Farmer | 58 | 21.0 |
|  | Daily Worker | 43 | 15.6 |
|  | Other | 45 | 16.3 |
| Smoking Status | Smoker | 135 | 48.9 |
|  | Non-Smoker | 141 | 51.1 |
| Alcohol | User | 163 | 59.1 |
|  | Non-User | 113 | 40.9 |
| Blood Pressure | Yes | 163 | 59.1 |
|  | No | 113 | 40.9 |
| Surgical History | Yes | 83 | 30.1 |
|  | No | 193 | 69.9 |
| Duration of Treatment | Short | 250 | 90.6 |
|  | Medium | 16 | 5.8 |
|  | Long | 10 | 3.6 |
| Type of treatment | Timolol | 99 | 35.9 |
|  | Pilocarpine | 26 | 9.4 |
|  | Timolol with Diamox | 13 | 4.7 |
|  | Timolol with Pilocarpine | 14 | 5.1 |
|  | Timolol with Diamox with Pilocarpine | 3 | 1.1 |
|  | Timolol with Dorzolamide | 29 | 10.5 |
|  | Dorzolamide with Latanoprost | 8 | 2.9 |
|  | Dorzolamide | 23 | 8.3 |
|  | Timolol with Latanoprost | 30 | 10.9 |
|  | OCT | 31 | 11.2 |
| Genetic History | Yes | 127 | 46.0 |
|  | No | 149 | 54.0 |

Regarding clinical and behavioural characteristics, 135 (48.9%) patients were smokers, 163 (59.1%) reported alcohol use, 163(59.1%) had a history of hypertension. On the other hand, 83(30.1%) had undergone glaucoma surgery, while 127 (46.0%) reported a family history of glaucoma. Regarding treatment characteristics, 250 (90.6)% glaucoma patients had a short term treatment duration, while 16(5.8%) had a medium treatment duration. Timolol was the most commonly used medication 99 (36.0%), followed by Timolol with Dorzolamide, Timolol with Latanoprost, and OCT, each accounting for 11.0%.

According to Table 2, the overall mean baseline intraocular pressure (IOP) was 17.31 (SD = 6.13) mmHg. At baseline, IOP was slightly higher in the left eye (OS: 17.52, SD = 7.64 mmHg) than in the right eye (OD: 17.11, SD = 7.16 mmHg). Some differences in mean IOP were observed across patient and clinical characteristics. Urban residents had a slightly higher mean IOP than rural residents (17.67 vs. 17.01 mmHg). Similarly, patients who reported alcohol use or smoking had somewhat higher mean IOP than those who did not (17.54 vs. 16.98 mmHg and 17.72 vs. 16.92 mmHg, respectively). In contrast, patients with a history of hypertension had a lower mean IOP than those without hypertension (16.78 vs. 18.08 mmHg). Mean IOP was broadly similar among patients with and without a history of surgical intervention (17.46 vs. 17.25 mmHg). The largest difference in baseline IOP was observed across treatment-duration groups. Patients with a medium treatment duration had the highest mean IOP (22.03 mmHg), with mean values of 20.12 mmHg in the right eye and 23.94 mmHg in the left eye. However, this group was relatively small (n = 16), and the relatively large standard deviations, particularly for the left eye, indicate substantial variability. Patients with short and long treatment durations had lower mean IOP values of 17.04 and 16.60 mmHg, respectively. Patients with a family history of glaucoma also had a slightly higher mean IOP than those without such a history (17.69 vs. 17.00 mmHg). Overall, these descriptive results suggest meaningful variation in baseline IOP across patient characteristics, although the observed differences should be interpreted cautiously because of the unequal and, in some cases, small subgroup sizes.

**Table 2.** Baseline Intraocular Pressure by Patient and Clinical Characteristics.

| Variables | categories | N | $IOP$ ($\bar{x}$ (sd)) | | $IOP_{OD}$ ($\bar{x}$ (sd)) | | $IOP_{OS}$ ($\bar{x}$ (sd)) | |
| --- | --- | --- | --- | --- | --- | --- | --- | --- |
| Residence | Rural | 148 | 17.01 | (6.65) | 16.59 | (7.35) | 17.42 | (8.06) |
|  | Urban | 128 | 17.67 | (5.07) | 17.7 | (6.91) | 17.64 | (7.17) |
| Alcohol use | Yes | 163 | 17.54 | (6.09) | 17.21 | (7.12) | 17.87 | (8.47) |
|  | No | 113 | 16.98 | (5.8) | 16.95 | (7.25) | 17.02 | (6.26) |
| Smoking | Yes | 135 | 17.72 | (5.79) | 17.57 | (6.95) | 17.87 | (8.27) |
|  | No | 141 | 16.92 | (6.13) | 16.66 | (7.35) | 17.18 | (7) |
| Blood Pressure | Yes | 163 | 16.78 | (5.23) | 16.59 | (6.42) | 16.97 | (6.05) |
|  | No | 113 | 18.08 | (6.85) | 17.85 | (8.08) | 18.32 | (9.46) |
| Surgical Intervention | Yes | 83 | 17.46 | (6.48) | 17.18 | (7.76) | 17.73 | (9.71) |
|  | No | 193 | 17.25 | (5.75) | 17.07 | (6.91) | 17.43 | (6.58) |
| Duration of Treatment | Short | 250 | 17.04 | (5.74) | 17.01 | (6.96) | 17.07 | (6.92) |
|  | Medium | 16 | 22.03 | (7.68) | 20.12 | (9.65) | 23.94 | (14.18) |
|  | Long | 10 | 16.6 | (6.02) | 14.6 | (6.88) | 18.6 | (6.74) |
| Genetically Hereditary | Yes | 127 | 17.69 | (5.94) | 17.66 | (7.66) | 17.71 | (7.82) |
|  | No | 149 | 17 | (6) | 16.63 | (6.69) | 17.36 | (7.51) |
| Average |  | 276 | 17.31 | (6.13) | 17.11 | (7.16) | 17.52 | (7.64) |

Figure 1 illustrates the longitudinal pattern of IOP and its baseline distribution across treatment groups and sex. As shown in Panel A, the mean IOP was consistently higher in the left-eye (OS) than that in the right-eye (OD) throughout the six follow-up visits. In the left eye, mean IOP increased slightly from baseline to the first follow-up visit and then gradually declined through visit 3, followed by a modest increase at visit 4 and another decline at visit 5. The right eye showed a clearer decline during the early follow-up visits, followed by a small increase at visit 4 and a subsequent decrease at visit 5.

**Fig. 1.**
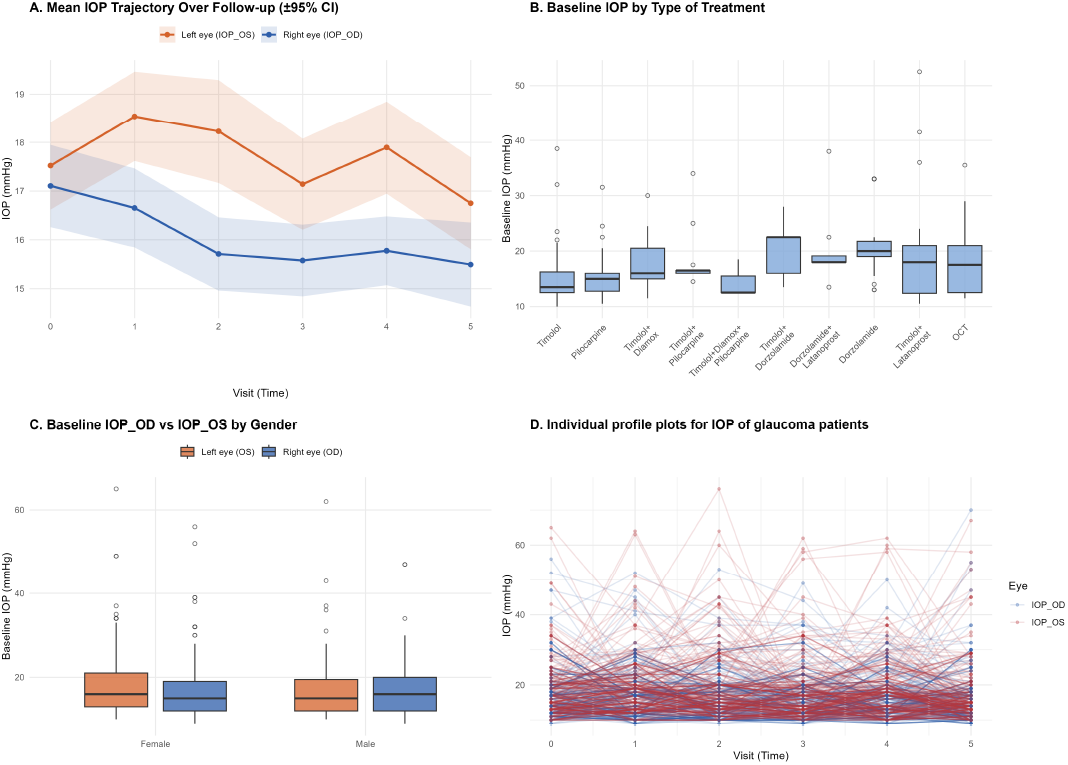
Longitudinal profile plot and Trajectories of Intraocular Pressure.

Panel B shows noticeable differences in the distribution of baseline IOP across treatment groups. Some groups had wider IOP distributions and several higher values, indicating substantial variation in baseline pressure among patients. However, these differences should be interpreted cautiously because some treatment groups contained relatively few patients. As shown in Panel C, the baseline distributions of IOP were broadly similar between males and females, although some variation between the two eyes was observed within each sex.

Panel D highlights the considerable variation in IOP trajectories among individual patients. While the average trajectories suggest an overall decline in IOP over follow-up, patients differed substantially in both their baseline IOP levels and the extent and direction of change over time. Overall, the figure indicates considerable between-patient variability and within-patient changes in IOP, together with a persistent difference between the two eyes. These patterns provide a clear rationale for using a longitudinal modeling approach that accounts for the correlation of repeated measurements within patients and differences in patient-specific trajectories.

According to Table 3, the 276 patients contributed a total of 1,656 IOP measurements over six follow-up visits. The mean age was 46.93 years (SD = 14.83), with ages ranging from 18 to 76 years. The mean IOP was higher in the left eye (OS) than in the right eye (OD), with mean values of 17.68 (SD = 8.10) and 16.05 (SD = 6.68) mmHg, respectively. The overall mean IOP was 16.87 (SD = 6.02) mmHg, with measurements ranging from 9.5 to 56 mmHg. The relatively large standard deviations indicate considerable variability in IOP measurements, particularly for the left eye. This was also reflected in the coefficients of variation, which were 45.8% for the left eye and 41.6% for the right eye, indicating greater relative variability in left-eye IOP measurements.

**Table 3.** descriptive statistics age and IOP across 1656 repeated measurements.

| Variable | N | Mean | SD | min | Max |
| --- | --- | --- | --- | --- | --- |
| Age | 1656 | 46.93 | 14.83 | 18 | 76 |
| Left eye (OS) | 1656 | 17.68 | 8.10 | 10 | 76 |
| Right eye (OD) | 1656 | 16.05 | 6.68 | 9 | 70 |
| IOP (average) | 1656 | 16.87 | 6.02 | 9.5 | 56 |

**Table 4.** Comparison of random-effects structures for longitudinal IOP data.

| **Right-eye IOP** |  |  |  |  |  |  |  |
| --- | --- | --- | --- | --- | --- | --- | --- |
| Random-effects structure | DF | AIC | BIC | Log-likelihood | LRT $\chi^2$ | LRT df | p-value |
| Random intercept | 28 | 9909.6 | 10061 | -4926.8 |  |  |  |
| Random intercept + slope | 30 | 9883.6 | 10046 | -4911.8 | 29.945 | 2 | 3.14e-07 |
| **Light-eye IOP** |  |  |  |  |  |  |  |
| Random intercept | 28 | 10325 | 10477 | -5134.5 |  |  |  |
| Random intercept + slope | 30 | 10317 | 10479 | -5128.5 | 11.951 | 2 | 0.0025 |
| **Bivariate IOP** |  |  |  |  |  |  |  |
| Random intercept | 30 | 20244 | 20427 | -10092 |  |  |  |
| Random intercept + slope | 33 | 20205 | 20406 | -10070 | 44.716 | 3 | 1.06e-09 |

Figure 2 displays the mean IOP trajectories according to residence and alcohol use across the six follow-up visits. The top panels present the mean IOP for the left-eye (OS) and right-eye (OD) by to residence. Overall, urban participants had higher mean IOP than rural participants throughout most of the follow-up period in both eyes. The difference was particularly evident for the left eye, while the mean right-eye IOP was similar between the two groups at the final visit.

**Fig. 2.**
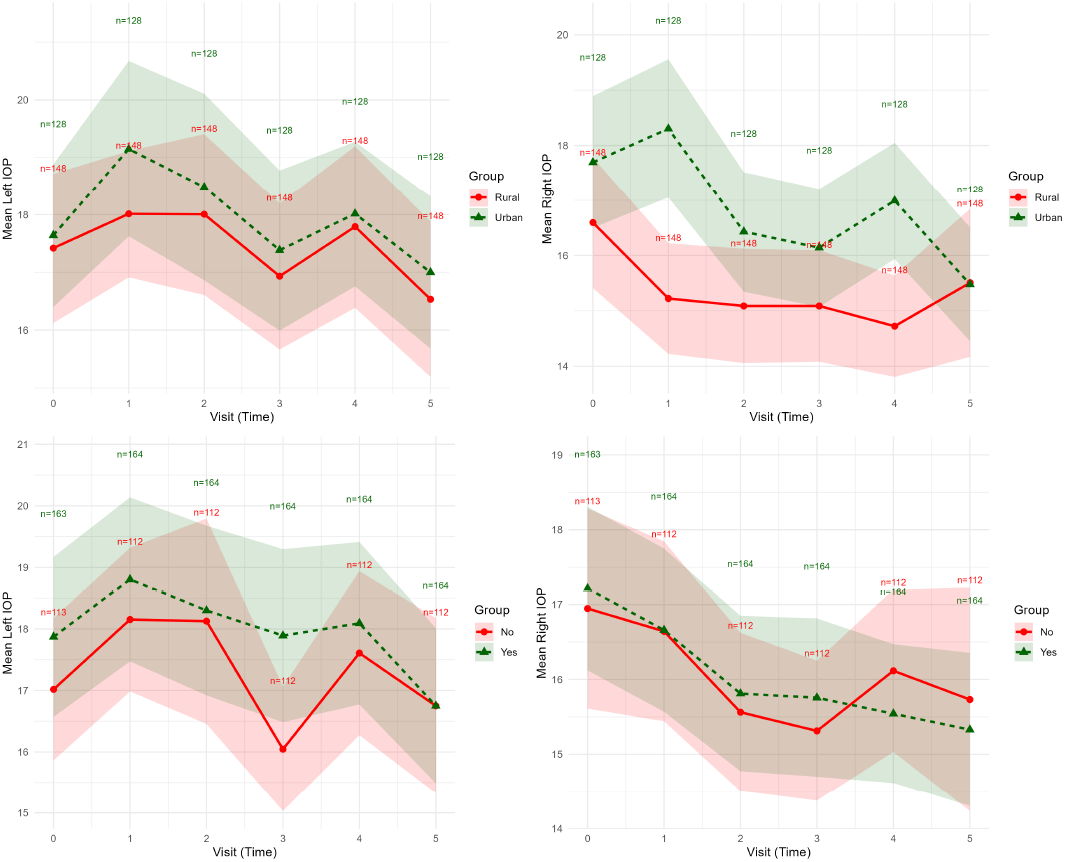
The mean profile plot of left- and right-eye IOP by residence on the top and by alcohol use on the bottom

The bottom panels present the mean IOP trajectories according to alcohol use. Alcohol users generally had higher mean IOP in the left eye than non-users across the follow-up visits. In the right eye, however, the pattern was less consistent, with alcohol users having slightly higher mean IOP during the earlier visits, whereas non-users had higher mean IOP at the fourth and sixth visits. These patterns suggest that differences in IOP according to alcohol use varied across follow-up visits and between the two eyes, whereas the pattern by residence was relatively more consistent.

### Linear Mixed Model Results

After evaluating the importance of fixed effects, the significance of random effects was assessed. This two-step approach allows for a thorough examination of both fixed and random effects in mixed-effects models. By first evaluating fixed effects and then incorporating random effects as needed, we can determine the best model that accounts for both the systematic and random sources of variability in the data, ensuring a robust and well-fitting final model for the analysis of IOP. To ensure valid inference from the linear mixed-effects models, the covariance structure of the repeated IOP measurements was appropriately specified. The random -slope model provided a significantly better fit than the random-intercept model for all left-eye, right-eye and bivariate IOP. The AIC values for the random-slope versus random-intercept models were 9883.6 vs. 9909.6, 10317 vs. 10325, 20205 vs. 20244 for right-eye IOP, left-eye IOP and bivariate IOP, respectively Table4. Thus, the random-slope model was selected for all three outcomes.

Table 5 presents the covariance structured considered for the repeated IOP measurements (left-eye, right eye and bivariate IOP) under the random intercept-slop model. Four commonly used covariance structures were evaluated: unstructured (UN),compound symmetry (CS), first-order autoregressive (AR(1)), and Toeplitz (TOEP). The unstructured covariance structure was selected for the final linear mixed-effects models because it yielded the smallest AIC and BIC values among the candidate covariance structures, indicating the best overall model fit.

**Table 5.** Comparison of covariance structure for linear mixed effect model.

| Covariance structure | AIC | BIC | Loglik |
| --- | --- | --- | --- |
| Left-eye (IOP_OS) |  |  |  |
| **Un** | **11282.79** | **11358.82** | **-5620.396** |
| TOEP | 11342.97 | 11364.69 | -5665.484 |
| AR(1) | 11459.32 | 11466.56 | -5727.662 |
| CS | 11379.84 | 11387.08 | -5687.920 |
| Right-eye (IOP_OD) |  |  |  |
| **Un** | **10655.48** | **10702.11** | **-5306.742** |
| TOEP | 10680.38 | 10731.51 | -5334.192 |
| AR(1) | 10822.83 | 10830.07 | -5409.414 |
| CS | 10757.34 | 10764.58 | -5376.668 |
| IOP |  |  |  |
| **Un** | **10276.86** | **10322.88** | **-5117.428** |
| TOEP | 10326.45 | 10348.17 | -5157.224 |
| AR(1) | 10462.57 | 10469.81 | -5229.287 |
| CS | 10396.61 | 10403.85 | -5196.307 |

Table 6 presents the linear mixed effect regression results for the separate left- and right-eye model and the bivariate model. The estimated coefficients represent differences in IOP relative to the corresponding reference categories, whereas the time-interaction terms represent differences in the rate of IOP change over follow-up. In the left-eye model, IOP decreased significantly over time male glaucoma patients had lower baseline IOP than females. Compared with civil servants, merchants and individuals in other occupations had significantly higher baseline IOP, whereas daily labourers had 0.13 mmHg lower baseline IOP (95% CI: *−* 0.25, *−* 0.004). Relative to Timolol, Timolol with Diamox, Timolol with Pilocarpine, Timolol with Xolamol, and Xolamol were associated with higher baseline IOP, with estimated increases of 0.185, 0.14, 0.131, and 0.18 mmHg, respectively. In contrast, Timolol with Latanoprost was associated with 0.12 mmHg lower baseline IOP. Short treatment duration was also associated with 0.129 mmHg lower baseline IOP compared with the reference duration. Significant time interactions were observed for selected treatment regimens and treatment duration. Compared with the reference occupation, the time-by-merchant interaction indicated a greater decline in IOP over time (*β* = *−* 0.029), although this association was borderline (*p* = 0.055). Relative to Timolol, the time-by-Timolol-with-Latanoprost interaction was associated with a slower decline in IOP over time (*β* = 0.038, *p* = 0.0197). Short treatment duration was also associated with a less pronounced decline in IOP over time (*β* = 0.0372, *p* = 0.0031).

**Table 6.**
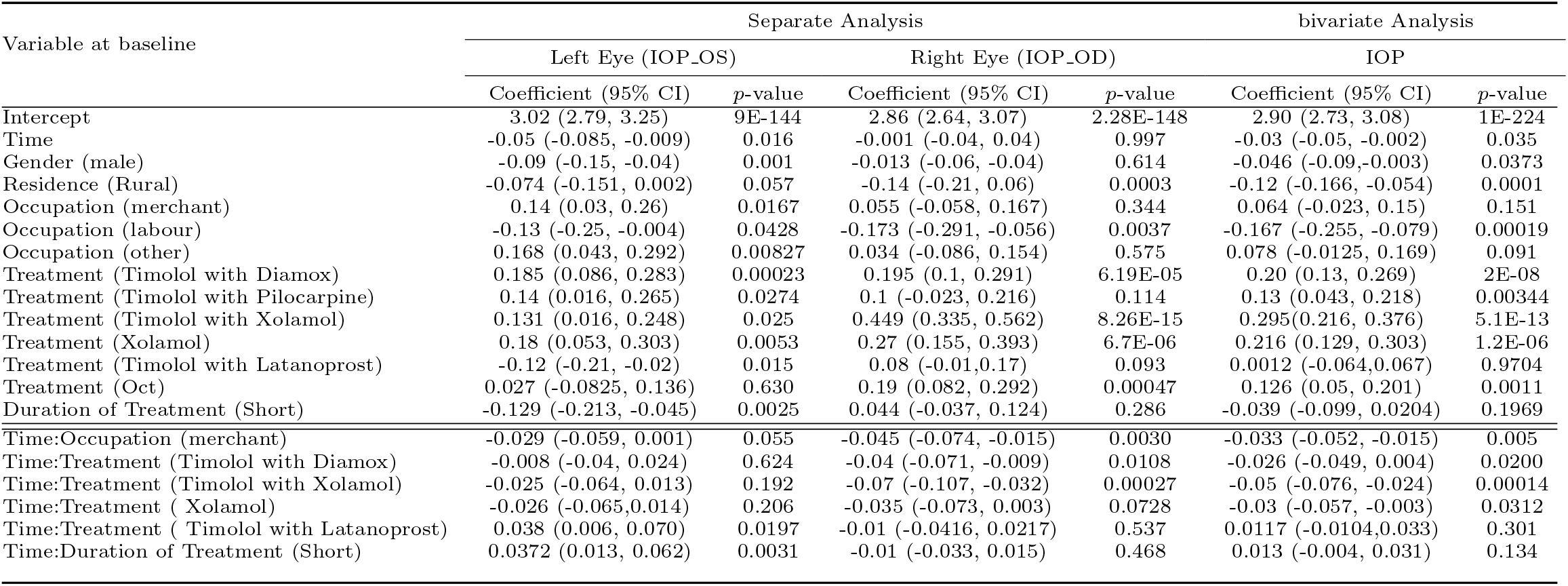
Regression results for separate and bivariate analyses of left- and right-eye (IOP_OS and IOP_OD)

| Variable at baseline | Separate Analysis |  |  |  | bivariate Analysis |  |
| --- | --- | --- | --- | --- | --- | --- |
|  | Left Eye (IOP_OS) |  | Right Eye (IOP_OD) |  | IOP |  |
|  | Coefficient (95% CI) | p-value | Coefficient (95% CI) | p-value | Coefficient (95% CI) | p-value |
| Intercept | 3.02 (2.79, 3.25) | 9E-144 | 2.86 (2.64, 3.07) | 2.28E-148 | 2.90 (2.73, 3.08) | 1E-224 |
| Time | -0.05 (-0.085, -0.009) | 0.016 | -0.001 (-0.04, 0.04) | 0.997 | -0.03 (-0.05, -0.002) | 0.035 |
| Gender (male) | -0.09 (-0.15, -0.04) | 0.001 | -0.013 (-0.06, -0.04) | 0.614 | -0.046 (-0.09, -0.003) | 0.0373 |
| Residence (Rural) | -0.074 (-0.151, 0.002) | 0.057 | -0.14 (-0.21, 0.06) | 0.0003 | -0.12 (-0.166, -0.054) | 0.0001 |
| Occupation (merchant) | 0.14 (0.03, 0.26) | 0.0167 | 0.055 (-0.058, 0.167) | 0.344 | 0.064 (-0.023, 0.15) | 0.151 |
| Occupation (labour) | -0.13 (-0.25, -0.004) | 0.0428 | -0.173 (-0.291, -0.056) | 0.0037 | -0.167 (-0.255, -0.079) | 0.00019 |
| Occupation (other) | 0.168 (0.043, 0.292) | 0.00827 | 0.034 (-0.086, 0.154) | 0.575 | 0.078 (-0.0125, 0.169) | 0.091 |
| Treatment (Timolol with Diamox) | 0.185 (0.086, 0.283) | 0.00023 | 0.195 (0.1, 0.291) | 6.19E-05 | 0.20 (0.13, 0.269) | 2E-08 |
| Treatment (Timolol with Pilocarpine) | 0.14 (0.016, 0.265) | 0.0274 | 0.1 (-0.023, 0.216) | 0.114 | 0.13 (0.043, 0.218) | 0.00344 |
| Treatment (Timolol with Xolamol) | 0.131 (0.016, 0.248) | 0.025 | 0.449 (0.335, 0.562) | 8.26E-15 | 0.295 (0.216, 0.376) | 5.1E-13 |
| Treatment (Xolamol) | 0.18 (0.053, 0.303) | 0.0053 | 0.27 (0.155, 0.393) | 6.7E-06 | 0.216 (0.129, 0.303) | 1.2E-06 |
| Treatment (Timolol with Latanoprost) | -0.12 (-0.21, -0.02) | 0.015 | 0.08 (-0.01, 0.17) | 0.093 | 0.0012 (-0.064, 0.067) | 0.9704 |
| Treatment (Oct) | 0.027 (-0.0825, 0.136) | 0.630 | 0.19 (0.082, 0.292) | 0.00047 | 0.126 (0.05, 0.201) | 0.0011 |
| Duration of Treatment (Short) | -0.129 (-0.213, -0.045) | 0.0025 | 0.044 (-0.037, 0.124) | 0.286 | -0.039 (-0.099, 0.0204) | 0.1969 |
| Time:Occupation (merchant) | -0.029 (-0.059, 0.001) | 0.055 | -0.045 (-0.074, -0.015) | 0.0030 | -0.033 (-0.052, -0.015) | 0.005 |
| Time:Treatment (Timolol with Diamox) | -0.008 (-0.04, 0.024) | 0.624 | -0.04 (-0.071, -0.009) | 0.0108 | -0.026 (-0.049, 0.004) | 0.0200 |
| Time:Treatment (Timolol with Xolamol) | -0.025 (-0.064, 0.013) | 0.192 | -0.07 (-0.107, -0.032) | 0.00027 | -0.05 (-0.076, -0.024) | 0.00014 |
| Time:Treatment (Xolamol) | -0.026 (-0.065, 0.014) | 0.206 | -0.035 (-0.073, 0.003) | 0.0728 | -0.03 (-0.057, -0.003) | 0.0312 |
| Time:Treatment (Timolol with Latanoprost) | 0.038 (0.006, 0.070) | 0.0197 | -0.01 (-0.0416, 0.0217) | 0.537 | 0.0117 (-0.0104, 0.033) | 0.301 |
| Time:Duration of Treatment (Short) | 0.0372 (0.013, 0.062) | 0.0031 | -0.01 (-0.033, 0.015) | 0.468 | 0.013 (-0.004, 0.031) | 0.134 |

In the right-eye model, rural residence was associated with 0.14 mmHg lower baseline IOP than urban residence (95% CI: *−* 0.21, 0.06). compared with civil servants, daily labourer had significantly lower baseline IOP (0.0173 mmHg, 95% CI: -0.291, -0.056). Relative to Timolol, Timolol with Diamox, Timolol with Xolamo, Xolamol and OCT were associated with higher baseline IOP, with estimated increases of 0.195, 0.449, 0.27 and 0.19 mmHg, respectively.

Significant time interactions were observed for merchant occupation and selected treatment regimens. Compared with the reference categories civil servant, merchants had a greater decline in IOP over time ( *β* = *−* 0.045 mmHg). Relative to Timolol, the time-by-treatment interactions for Timolol with Diamox and Timolol with Xolamol were associated with additional declines in IOP of 0.04 and 0.035 mmHg per unit increase in time, respectively.

In the bivariate model, rural residence was associated with 0.12 mmHg lower baseline IOP than urban residence (95% CI: *−* 0.166, *−* 0.054). Compared with civil servants, daily labourers had significantly lower baseline IOP by 0.167 mmHg (95% CI: *−* 0.255, *−* 0.079). Relative to Timolol, Timolol with Diamox, Timolol with Pilocarpine, Timolol with Xolamol, Xolamol, and OCT were associated with higher baseline IOP, with estimated increases of 0.20, 0.13, 0.295, 0.216, and 0.126 mmHg, respectively. The time-by-merchant interaction indicated a greater decline in IOP over time (*β* = *−* 0.033, *p* = 0.005). Significant time-by-treatment interactions were also observed for Timolol with Diamox, Timolol with Xolamol, and Xolamol, corresponding to additional declines of 0.026, 0.05, and 0.03 mmHg per unit increase in time, respectively.

Overall, the linear mixed-effects analysis showed that IOP varied both between patients and over time. Residence, occupation, treatment regimen, and treatment duration were associated with baseline IOP, although the strength and direction of these associations differed between the left and right eyes. Several treatment regimens were also associated with different rates of IOP change during follow-up, suggesting that the longitudinal response to treatment was not uniform across patients. The bivariate analysis generally supported the main findings from the separate eye-specific models while accounting for the correlation between the two eyes. These findings highlight the importance of considering both patient-level differences and the longitudinal nature of IOP measurements when assessing glaucoma outcomes.

## 4 Discussion

The objective of this study was to determine risk-factors that associated with the progression of intraocular pressure of glaucoma patients by using linear mixed effect model. IOP is known as an important factor for glaucoma pathogenesis [20]. From the results of linear mixed model, age is statistically non-significant. Several studies have shown that IOP increased with age, whereas others suggested no relationship with age. IOP changed during follow-up, although the pattern differed between the two eyes. Left-eye IOP decreased significantly over time, whereas no significant effect over the right-eye IOP change. Similar reductions in IOP over follow-up have been reported among glaucoma patients in Felege Hiwot Referral Hospital, Ethiopia [21, 22]. The difference between the two eyes in our study may reflect variation in disease severity, treatment response, or other eye-specific factors. In a Barbados Eye Study the mean IOP increased with 1 mmHg for every 10 years of age increment [23]. However, a Japanese study showed that IOP correlated inversely with age in men and women [24]. Gender also appears to have an effect on IOP [7]. Studies have shown that there may be differences in the prevalence of glaucoma between men and women, with some types of glaucoma being more common in one gender over the other [25]. Hormonal changes, genetic factors, and differences in eye anatomy between genders may contribute to these variations in glaucoma prevalence [26]. In this study Gender was significant in the left-eye IOP and the percentage of males were higher than females.

According to this study residence was significantly associated with baseline IOP, with rural glaucoma patients having lower IOP than urban patients in the right-eye and bivariate model. An Ethiopian study also reported a significant association between residence and IOP, although the direction of the association differed from our findings [21]. Our findings show that urban dwellers had greater IOP compared with Rural which is consistent with the study [27]. Such differences may be related to variations in access to eye-care services, treatment patterns, disease severity, and socioeconomic characteristics between study populations. Therefore, residence may be acting as a marker of differences in healthcare access and patient characteristics rather than having a direct effect on IOP.

Occupation was also associated with baseline IOP in the present study. Daily labourer had lower baseline IOP than than civil servants, while occupation was associated with the rate of change in IOP over time in the right-eye model [28]. This finding is consistent with previous Ethiopian evidence in which occupation was associated with IOP among glaucoma patients [22]. The observed differences may reflect socioeconomic circumstances, treatment adherence, access to healthcare, or other factors associated with occupation rather than a direct effect of occupation itself. However, further research is needed to clarify the mechanisms underlying this association.

Treatment regimen was strongly associated with baseline IOP in our study, and several treatment groups also showed different rates of change over time. In the left-eye model, Timolol with Diamox, Timolol with Pilocarpine, Timolol with Xolamol, Xolamol and Timolol with Latanoprost were significantly associated with IOP, ndicating differences in IOP trajectories across treatment regimens [29]. Similarly, Timolol with Diamox, Timolol with Xolamol, Xolamol, and OCT were significantly associated with IOP in the right-eye model. These findings highlight the importance of treatment regimen in the longitudinal management of IOP among patients with glaucoma, although the observed associations should not be interpreted as evidence that one regimen is more effective than another, given the observational nature of the study [30]. Evidence from several sub-Saharan African countries also shows considerable variation in glaucoma treatment patterns, reflecting differences in disease severity, clinical practice, and availability of treatment [31].

The higher IOP observed in the left eye than in the right eye is also consistent with previous observations among Ethiopian glaucoma patients, although the magnitude of the difference varies between studies. More importantly, maintaining adequate IOP control is clinically relevant for limiting glaucoma progression. An Ethiopian multicentre study found that patients who remained stable over five years had substantially lower mean IOP than those who progressed, highlighting the importance of sustained IOP reduction during long-term management [32].

According to this study, Duration of treatment is had been found a significant effect for the progression of left-eye IOP of glaucoma patients. Duration of treatment (long time) has a significant negative effect on the response variable [33, 34]. When we come to the joint analysis, residence, occupation, treatment type and duration of treatment have a significant effect on IOP [22].

The longitudinal mixed effect analysis over a follow-up differed according to selected baseline characteristics, particularly occupation, treatment type, and treatment duration. Time with occupation(merchant) interaction in the right eye and joint analysis suggests that merchants experienced a greater decline in IOP over time. Treatment related interactions were more consistent. The treatment regime not only influence the baseline level of IOP but also its subsequent trajectory in particular time interaction with (Timolol with Diamox) significantly decline 0.85 mmHg per time unit for the right-eye IOP and 0.57 mmHg in the joint analysis. Similarly, Timolol with Xolamol was associated with an additional decline of 1.41 mmHg per time unit in the right eye and 1.00 mmHg in the joint analysis. These findings is consistent with the previous studies suggested that long term follow-up sustained reduction with Timolol-based combinations [35, 36].

There is also a significant greater decline in IOP over time among patients with long treatment duration for the left eye and in the joint analysis. In general, the rate of IOP changes depends on treatment type, occupation and duration of treatment. The stronger treatment-related change observed in the right-eye suggested that the importance of considering the two eyes separate analysis rather than averaging IOP measure [37].

Overall, our findings are broadly consistent with previous Ethiopian and sub-Saharan African evidence showing that IOP trajectories are influenced by follow-up time, treatment, residence, occupation, and patient characteristics. By jointly modelling the two eyes while also examining them separately, this study provides a more complete picture of longitudinal IOP variation. These findings support individualized monitoring of glaucoma patients and emphasize the importance of considering changes in IOP over time when evaluating treatment and disease management.

## 5 Conclusions

The linear mixed-effects model (LMM) identified Gender, residence, occupation, treatment regimen, and duration of treatment as significant predictors of longitudinal intraocular pressure (IOP). Treatment regimen was significantly associated with changes in IOP over time, indicating that IOP trajectories varied across treatment groups. Some treatment regimens, particularly Timolol with Diamox, Timolol with Xolamol, and Xolamol, were associated with greater reductions in IOP over time. The mean IOP was consistently higher among patients residing in urban areas than among those residing in rural areas. Shorter treatment duration was also associated with a greater reduction in mean IOP. Differences in baseline disease severity, treatment selection, adherence, and clinical management may have contributed to the observed longitudinal patterns. Overall, these findings highlight the importance of demographic and clinical factors, particularly residence, occupation, treatment regimen, and treatment duration, in understanding longitudinal patterns of IOP among glaucoma patients.

## Data Availability

For Study Protocols: No datasets were generated or analysed during the current study. All relevant data from this study will be made available upon study completion.

